# AI-Detected Asymptomatic Atrial Fibrillation and Risk of Incident Ischemic Stroke and Cardiovascular Events: A UK Biobank Study

**DOI:** 10.64898/2026.02.13.26346138

**Authors:** Arjun K. Butani, Zareen Farukhi, David Brüggemann, Pamela Rist, Felix C. Tanner, Olga V. Demler

## Abstract

**Background:** Advances in wearable devices and machine-learning-based ECG analysis enable highly accurate detection of atrial fibrillation (AF) outside traditional clinical settings, leading to increasing identification of asymptomatic AF. However, the prognostic significance of AI-detected asymptomatic AF and its implications for downstream cardiovascular risk remain unclear. In contrast to clinically diagnosed AF, evidence guiding risk stratification and further evaluation in this population is limited. We therefore investigated the association between AI-detected asymptomatic AF and incident cardiovascular outcomes in a large population-based cohort.

**Methods:** We applied a validated open-source ECG-based deep learning model for atrial fibrillation detection (AI-AF) to 12-lead ECG recordings from participants in the UK Biobank. Participants with AI-detected AF on ECG and no prior clinical AF diagnosis were classified as asymptomatic AF (c). Kaplan-Meier curves and log-rank tests were used to compare the incidence of ischemic stroke and major adverse cardiovascular events (MACE: myocardial infarction, ischemic stroke, or cardiovascular death) across AF subgroups. Cox proportional hazards models were used to evaluate the association between AI-AF risk and incident MACE, adjusting for age, sex, current smoking, systolic blood pressure, total and HDL cholesterol, and prevalent type 2 diabetes. Follow-up was administratively censored at 6 years.

**Results:** The study included 96,531 participants with mean [SD] age of 65 [8] years; 52% female; median follow-up [IQR] of 4.7 [1.6-7.2] years. ECG data were available for 64,029 participants and an additional 32,502 participants with clinically diagnosed atrial fibrillation (AF) without ECG recordings were included. Among participants without prior clinical AF and with available ECGs, 2,399 were classified as asympAF based on AI detection, while 58,879 were AF-free. Over 6 years of follow-up, the incidence of ischemic stroke was significantly higher in participants with asympAF compared with AF-free individuals (1.5% vs 0.52%, p = 7×10^−7^) and significantly lower than in participants with clinically diagnosed AF (1.5% vs 3.4%, p = 2×10^−5^). Similar patterns were observed for myocardial infarction and cardiovascular death. Using a more liberal AI-AF threshold corresponding to a 15% false-positive rate (asympAF_15_) yielded consistent findings: participants classified as asympAF_15_ had a 62% higher risk of incident MACE in adjusted Cox PH models (hazard ratio 1.6, 95% CI 1.2-2.2) over six years.

**Conclusion:** AI-detected asymptomatic AF identified individuals at elevated risk of ischemic stroke and major adverse cardiovascular events. As ischemic stroke is a hallmark complication of atrial fibrillation, these findings support the hypothesis that AI-ECG models may capture subclinical AF-related risk not detected by conventional clinical assessment. This approach may help extend the window for preventive interventions in populations without clinically diagnosed AF.

## Introduction

In the recent past, clinically diagnosed atrial fibrillation (AF) typically was based on ECG images recorded in clinician office for patients who have symptoms such as palpitations, chest pain, dizziness, fatigue, lightheadedness, reduced ability to exercise and/or shortness of breath. With recent advances in wearable technology and deep learning, ECGs from wearable devices have become commonplace, enabling the detection of AF through smartwatches and similar devices. Accuracy of AF detected from wearable devices is high (AUC about.98) and FDA has approved use of AF diagnosis read from wearable devices using machine learning models for research purposes^1^. AF readings from wearable devices made AF diagnosis readily available for millions of consumers which led to a multiple fold increase in patient visits for asymptomatic AF (asympAF). The standard follow-up for clinically-diagnosed AF averages thousands of dollars per patient. Given the surge in asymptomatic AF detection with wearable devices, the benefit of such extensive follow-up for asymptomatic AF is unknown. In this manuscript we used an open-source deep learning model to process ECG images from publicly available ECG repositories and 64,029 patients from UK Biobank to split into clinicalAF, asympAF and no AF groups. We compared the risk of developing future cardiovascular conditions for participants with asympAF versus clinicalAF versus AF-free individuals using Cox regression models adjusted for cardiovascular risk factors.

## Methodology

### Dataset

Data was derived from the UK Biobank (UKB), a large-scale prospective cohort study comprising over 500,000 participants between the ages of 40 and 69 years with detailed phenotypic, genetic, and imaging data. Out of 66,031 individuals with available 12-lead ECG recordings, 64,029 had ECG that passed quality control and were included in the analysis. Recordings with missing or unreadable data, as well as those from participants who had experienced cardiovascular events prior to the ECG, were excluded. The baseline for participants with ECG was set to the date of the ECG recording. This approach was used to avoid immortal time bias in comparisons between clinically diagnosed AF and AI-detected AF groups. All participants with clinically-diagnosed AF were included regardless of whether they had an ECG recording In UK Biobank. In this case, to avoid immortal bias, their baseline was set to the date of their index AF diagnosis. With the addition of this criterion, the total sample size consisted of 96,531 participants.

### Model Used and its Validation

A pretrained convolutional neural network model developed by Ribeiro et al.^2^ was used for AF detection. The model’s performance was validated using two independent ECG datasets, PTB-XL^3-5^ and CPSC 2018^6^, demonstrating high classification accuracy with an AUC of 0.99 and 0.94 for AF detection, respectively.

### Signal Processing

To optimize the signal quality for model application, the ECG signals were processed using a high-pass Butterworth filter with a cut-off frequency of 0.75 Hz and a low-pass Butterworth filter at 50 Hz to remove baseline drift and high-frequency noise. The UKB ECGs were pre-filtered to remove 50/60 Hz powerline interference. The signals were resampled to 400 Hz and the voltage units were standardized to 10^-4^V as required by the deep learning model. To capture as many AF cases as possible, the selection threshold was lowered such that the false positive rate (FPR) was 15% with the goal to maximize the true positive rate (TPR). The resulting TPR was 99% for PTB-XL.

### Classification Definition

The participants were separated into 3 different groups, asymptomatic atrial fibrillation (asympAF), clinically-diagnosed atrial fibrillation (clinicalAF), and no AF. If a participant was identified with AF using the AI model’s lenient threshold but had no prior clinical diagnosis, they were classified as asympAF. Those with a pre-existing clinical AF diagnosis were classified as clinicalAF.

### Outcome Definition

The primary outcome analyzed was a composite of ischemic stroke and major adverse cardiovascular event (MACE: IS, MI and CV death).

### Statistical Analysis

Kaplan-Meier survival curves were generated. Log-rank tests were used to test the differences in the incidence of the outcome between subgroups using type I error of.05. We reported the p-value and cumulative incidence of events at 6 years after the index date, as follow-up for the majority of participants did not extend beyond this time point. Cox proportional hazards (Cox PH) models were used to adjust for known confounders, including age, sex, smoking status, systolic blood pressure, cholesterol levels, and diabetes status. SCORE2, a validated cardiovascular risk model, was computed for all participants to contextualize AI-detected AF within established risk stratification frameworks^7^. Therefore, SCORE2 served as a benchmark to compare the predictive value of AI-detected asymptomatic AF with established clinical risk stratification models.

## Results

### Study cohort

The study included a total of 96,534 participants. In UK Biobank, 35,253 had a clinical diagnosis of symptomatic AF. Out of those without clinical AF and with ECG recording available, 2,399 were identified with asymptomatic AF using the AI model, and remaining 58,789 as free from AF (Figure).

**Figure.**
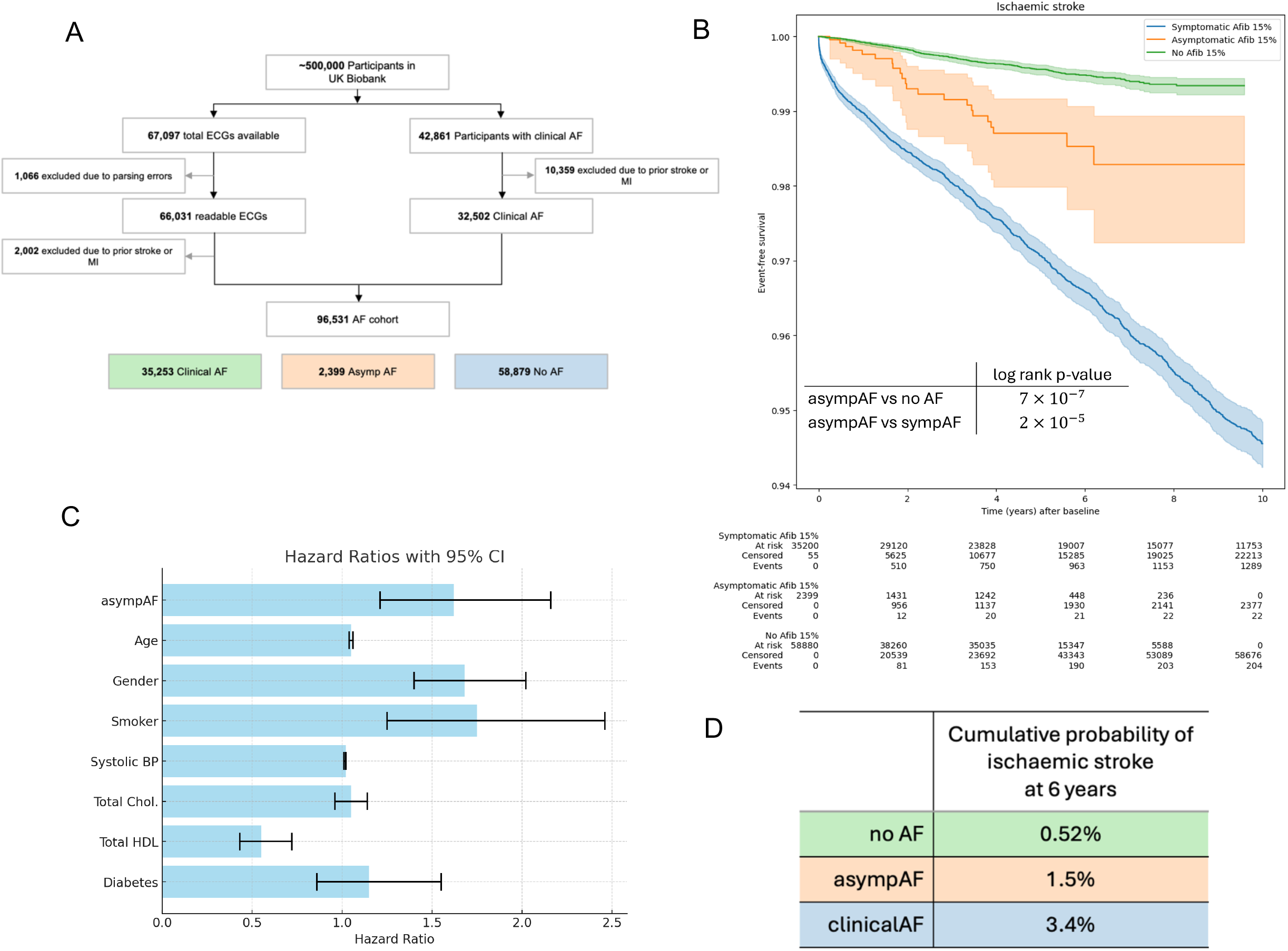
A: Consort diagram of study design. B: Kaplan Meier Curve of incident ischemic stroke among symptomatic AF (blue), asymptomatic AF (orange), no AF (green) in UK Biobank; C: Performance of asymptomatic AI-ECG AF diagnosis in predicting incident MACE: Cox proportional hazard regression model adjusted for age, gender, smoking (ever), systolic blood pressure, total and HDL cholesterol and type 2 diabetes; D: Cumulative incidence of ischaemic stroke at 6 years after index ECG.

The mean age in the cohort was 65.8 years (SD: 8.5). The asympAF group was on average older than the clinicalAF and the no AF groups (69.1 vs 67.2 vs 64.9, p < 0.001), had a higher proportion of males (73.3% vs 60.2% vs 45.4%, p < 0.001) and a higher mean systemic coronary risk evaluated using SCORE2 (11.1 vs 9.5 vs 7.5, p < 0.001). The AF group (asympAF + clinicalAF) had higher mean systolic blood pressure when compared with the no AF group (142.6 vs 139.7 mmHg, p < 0.001). It should be noted that the cholesterol data had a lot of missing entries. Table 1 describes the distribution of the participants in more detail.

**Table 1.**
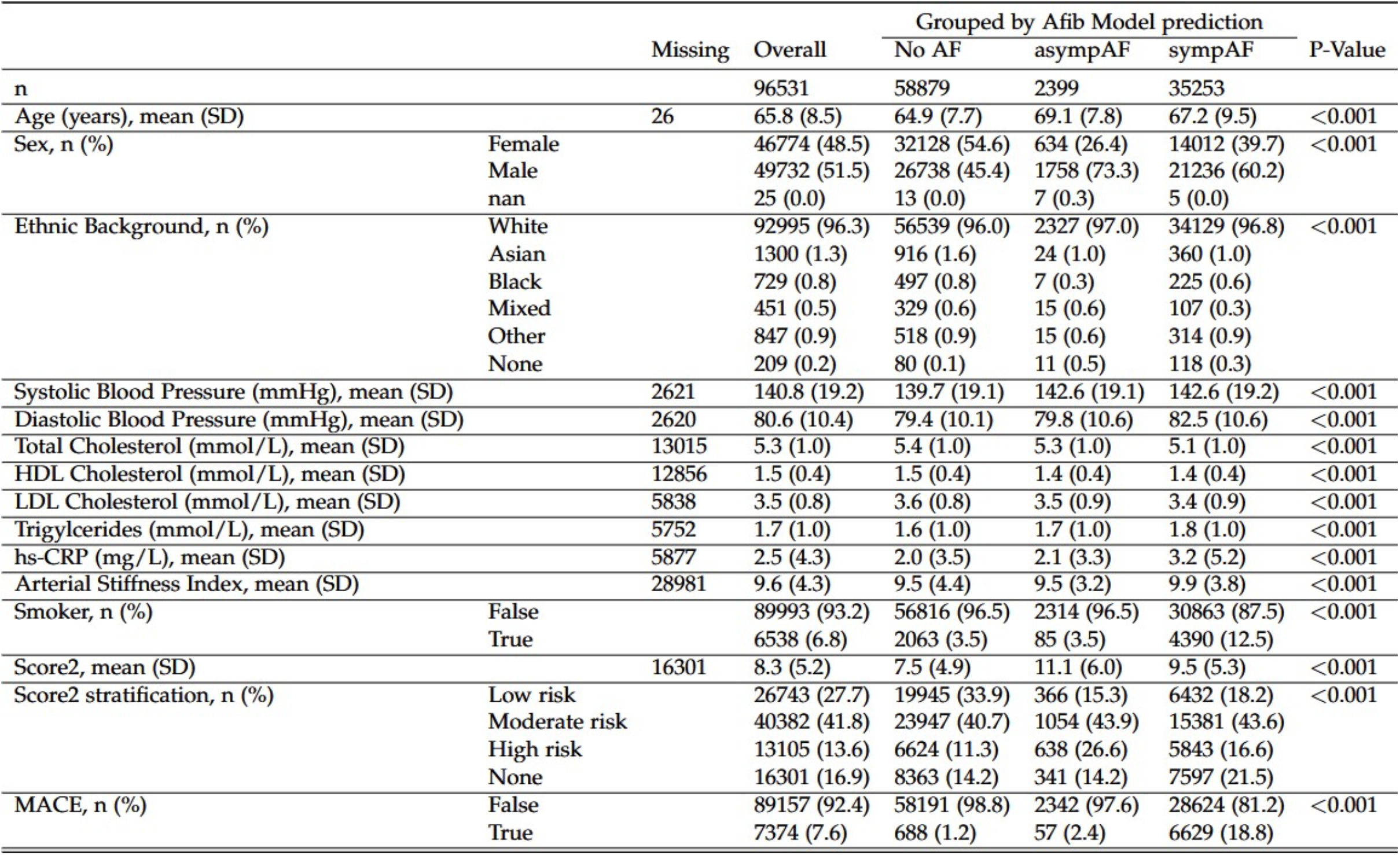
Demographic and clinical characteristics of the study cohort in the UK Biobank.

### Kaplan-Meier and Cox Proportional Hazards Regression Model

During the median follow-up period of 4.7 years (IQR: [1.6, 7.2] years) which was administratively censored at 6 years, there were 1,174 cases of ischemic stroke, 1,958 MI and 2,423 cardiovascular death events. The incidence of ischaemic stroke (IS) differed significantly across groups. Participants with asympAF had a significantly higher incidence of IS compared to those without AF (1.5% vs 0.52%, p =7·10^-7^). ClinicalAF participants had a higher risk of IS than asympAF (3.4% vs 1.5%, p = 2·10^-5^) (Figure). Similar results were observed for myocardial infarction, CVD death and for composite endpoint of MACE. Participants with asympAF exhibited a higher likelihood for cardiovascular death compared to those without AF (1.57% vs 0.41%, p = 3·10^-6^). Similar to the IS, there was a significant difference in the cardiovascular death incidences between participants with asympAF versus those with clinicalAF (1.57% vs 7.57%, p = 4·10^-22^). In adjusted Cox proportional hazards (Cox PH) regression analysis participants with asymptomatic AF had higher hazard ratio of developing MACE (HR 1.62; 95% CI [1.2, 2.2], p<0.001) than no AF participants (Figure).

### SCORE2

For the participants who experienced MACE 33 out of 57 participants with detected asympAF were categorized as moderate risk and 2 out of 57 as low risk by SCORE2. The discrepancy between AI-based and SCORE2 predictions highlights that there may be potential advantages to enhancing the SCORE2 predictor by assigning higher risk to participants who were diagnosed with asympAF by the AI model.

### Limitations

Definition of symptomatic/asymptomatic atrial fibrillation depends on quality of medical history recording. The first occurrence of clinically diagnosed AF in UK Biobank is recorded from the primary care data, the hospital inpatient data, death register records, and self-reported medical condition codes. The former two rely on centrally collected data from the national registry. Asymptomatic AF was defined based on ECGs of patients without clinical AF diagnosis. This setting is close to accidental finding of AF from a wearable device. Another limitation is whether AI-ECG model quality^2^ is comparable to that coming from a wearable device. Given that FDA approved AF from wearable devices for research purposes, the two settings are likely relatively close in performance.

Additionally, UK Biobank is an extensive dataset, but its scope is limited to the UK population, which may introduce some regional bias into the data. This limitation raises concerns about the generalizability of the findings derived from the UKB to other populations or global contexts. This can be addressed by comparing the results of this study with a similar analysis on a different dataset comprising participants from a different region or even a global dataset.

Finally, further investigations are needed to extend the results of this study to single-lead ECGs from wearable devices. This would help to establish whether early changes in ECG signal extend the window of opportunity for effective preventative treatments that can stop or reverse development of AF and ultimately prevent future cardiovascular events.

## Conclusions

This study demonstrates that AI-detected AF is associated with a significantly elevated risk of major adverse cardiovascular events. The findings suggest that deep learning models applied to ECG data can serve as a valuable tool for early risk stratification, identifying individuals who may benefit from closer monitoring or preventive interventions. Given the increasing use of wearable and AI-powered ECG devices, further research is needed to explore the clinical utility of AI-detected AF in guiding therapeutic decision-making. AI-based AF detection identified high-risk individuals who were not captured by the SCORE2 risk assessment, suggesting that traditional cardiovascular risk models may not capture the same information as the AI-detected AF models. Future studies should aim to replicate these results and assess the impact of early intervention strategies in asymptomatic individuals identified as high risk by AI models.

## Data Availability

All data produced in the present study are available upon reasonable request to the authors. UK Biobank data is available conditional on institutional approval between UK Biobank and third party.

https://www.ukbiobank.ac.uk/

## Acknowledgements

This research has been conducted using the UK Biobank Resource under application number 81959 and 206575.

## IRB approval numbers

BASEC-Nr.: Req-2024-00223; IRB protocol# 2020P002478.

